# Perceptual and semantic deficits in face recognition in semantic dementia

**DOI:** 10.1101/2024.07.10.24310157

**Authors:** Golnaz Yadollahikhales, Maria Luisa Mandelli, Zoe Ezzes, Janhavi Pillai, Buddhika Ratnasiri, David Paul Baquirin, Zachary Miller, Jessica de Leon, Boon Lead Tee, William Seeley, Howard Rosen, Bruce Miller, Joel Kramer, Virginia Sturm, Maria Luisa Gorno-Tempini, Maxime Montembeault

## Abstract

**State of the art:** Semantic dementia (SD) patients including semantic variant primary progressive aphasia (svPPA) and semantic behavioral variant frontotemporal dementia (sbvFTD) patients show semantic difficulties identifying faces and known people related to right anterior temporal lobe (ATL) atrophy. However, it remains unclear whether they also have perceptual deficits in face recognition.

**Methodology:** We selected 74 SD patients (54 with svPPA and predominant left ATL atrophy and 20 with sbvFTD and predominant right ATL atrophy) and 36 cognitively healthy controls (HC) from UCSF Memory and Aging Center. They underwent a perceptual face processing test (Benton facial recognition test-short version; BFRT-S), and semantic face processing tests (UCSF Famous people battery – Recognition, Naming, Semantic associations – pictures and words subtests), as well as structural magnetic resonance imaging (MRI). Neural correlates with the task’s performance were conducted with a Voxel-based morphometry approach using CAT12.

**Results:** svPPA and sbvFTD patients were impaired on all semantic face processing tests, with sbvFTD patients performing significantly lower on the famous faces’ recognition task in comparison to svPPA, and svPPA performing significantly lower on the naming task in comparison to sbvFTD. These tasks predominantly correlated with gray matter (GM) volumes in the right and left ATL, respectively. Compared to HC, both svPPA and sbvFTD patients showed preserved performance on the perceptual face processing test (BFRT-S), and performance on the BFRT-S negatively correlated with GM volume in the right posterior superior temporal sulcus (pSTS).

**Conclusion:** Our results suggest that early in the disease, with the atrophy mostly restricted to the anterior temporal regions, SD patients do not present with perceptual deficits. However, more severe SD cases with atrophy in right posterior temporal regions might show lower performance on face perception tests, in addition to the semantic face processing deficits. Early sparing of face perceptual deficits in SD patients, regardless of hemispheric lateralization, furthers our understanding of clinical phenomenology and therapeutical approaches of this complex disease.

## 1. Introduction

Semantic dementia (SD) is one of the main clinical variants of frontotemporal dementia (Hodges et al., 2007). Progressive ATL atrophy as a hallmark of the disease pathophysiology can lead to two different syndromes depending on the predominant side of atrophy. On one hand, svPPA patients is characterized by left-predominant ATL atrophy as well as by anomia and single-word comprehension deficits (M. L. Gorno-Tempini et al., 2011). On the other hand, sbvFTD patients with predominantly right sided ATL atrophy have profound changes in emotion and behavior, which can be hard to distinguish from those of the behavioral variant of frontotemporal dementia (bvFTD) and psychiatric disorders. While consensus criteria for sbvFTD are not yet available, it was recently proposed that loss of empathy (difficulty understanding emotions), complex compulsions or rigid thought process and difficulty naming and identifying known people are the core features of this clinical syndrome (Younes et al., 2022). The famous face recognition deficits observed in the sbvFTD patients are of particular interest, as they may be the most specific symptom of sbvFTD.

Previous studies have shed some light on the face recognition deficits seen in SD. Famous people have been commonly utilized as stimuli due to their integral role in shared semantic knowledge within specific cultures. Research findings indicate that patients with SD, irrespective of the predominant side of ATL atrophy, encounter difficulties in naming famous people and providing semantic information about them, such as their occupations or reasons for their fame (Binney et al., 2016; Ding et al., 2020; Irish et al., 2013; Kamminga et al., 2015; Luzzi et al., 2017; Mendez et al., 2015; Montembeault et al., 2017; Pozueta et al., 2019; Snowden et al., 2012; Younes et al., 2022). Some authors have proposed that these challenges are more pronounced in SD patients with a predominant left ATL atrophy, attributing it to the verbal nature of the task which necessitates lexical access (Snowden et al., 2012). However, numerous studies have demonstrated no between-group differences(Binney et al., 2016; Ding et al., 2020; Luzzi et al., 2017; Younes et al., 2022) using naming tasks, or even more pronounced deficits in SD patients with a predominant right ATL atrophy(Irish et al., 2013; Mendez et al., 2015).Other researchers have employed tasks specifically designed to isolate semantic abilities, eliminating the need for verbal responses that involve lexical access. Such tasks encompass matching a famous person’s name with the corresponding picture from a set of distractors or utilizing a triplet task where participants are tasked with matching a picture of a famous person with another picture of someone in the same field of celebrity, amid distractors. Similarly, patients with SD are consistently impaired on these types of tasks, with studies suggesting equivalent or more severe impairment in patients with right-predominant ATL atrophy, in comparison to those with left-predominant ATL atrophy(Ding et al., 2020; Luzzi et al., 2017; Younes et al., 2022). Perhaps most interestingly, famous face familiarity tasks, which involve selecting the image of a famous person amongst many distractors, appears to be the most specific impairment in sbvFTD. In contrast, svPPA patients show preserved performance in this type of task(Binney et al., 2016; Younes et al., 2022).

Although deficits in face recognition have been consistently demonstrated in SD, their underlying cognitive mechanisms remain partially elucidated. Models of face processing suggest a perceptual stage which includes early perception of facial features followed by extraction of invariant facial features, while simultaneously accounting for any changeable aspects of faces such as lip movements during speech. The next step in the process of face recognition is the semantic stage which results in the retrieval of biographical/semantic information related to the face. On one hand, it appears clear that the core semantic impairment in SD affect their ability to recognize known people. This impairment extends beyond the use of faces as stimuli and is genuinely multimodal, as observed in difficulties with recognizing famous people’s voices and names(Borghesani et al., 2019; Luzzi et al., 2017). On the other hand, the perceptual nature of these deficits has been rarely investigated, yielding mixed results (Chen et al., 2018; Ding et al., 2020; Kamminga et al., 2015; Kumfor et al., 2015). It is therefore crucial to establish whether the inability to recognize familiar faces seen in SD patients is merely related to a semantic deficit or if it is also due to a deficit at the face perception stage (Gainotti, 2007; Joubert et al., 2006; Kamminga J, 2015).

Moreover, the face recognition network is subdivided into a ‘core’ system which is responsible for primarily perceptual processing and an ‘extended’ network that underpins cognitive aspects of processing, including accessing person knowledge and making inferences about the person’s state and intentions (Haxby et al., 2000). Posterior occipital and temporal regions, such as the fusiform face area (FFA), the occipital face area (OFA), and the posterior superior temporal sulcus (pSTS), appear to comprise a core face processing system (Gobbini et al., 2007; Natu et al., 2011). Additionally, proceeding along a posterior-to-anterior axis, responses become increasingly tuned to more complex feature combinations, ultimately ending with higher-order semantic processing within the ATL (Binney et al., 2012; Brambati et al., 2010). Establishing the neural correlates of semantic and perceptual face processing in SD could offer further validation to the existing model, utilizing SD as a lesion model. Additionally, it has the potential to elucidate the localization of face processing symptoms within this population.

In the present study, we first investigated the performance of a large sample of HC and SD patients subdivided in svPPA and sbvFTD on perceptual and semantic face processing tasks. Second, we used voxel-based morphometry (VBM) to identify the neural correlates of perceptual and semantic face processing tasks.

## 2. Methods

### 2.1 Participants

Patients were recruited through the University of California San Francisco (UCSF) Memory and Aging Center (MAC) between 2003 and 2022. Patients with svPPA fulfilled the current diagnostic criteria for imaging-supported svPPA (1), while patients with sbvFTD fulfilled the proposed criteria for probable sbvFTD suggested by Younes and colleagues (2). A comprehensive evaluation (neurological history and examination, standardized neuropsychological and language evaluations) and a review of this evaluation at a consensus diagnostic meeting at the UCSF MAC were used to make the diagnosis. The predominance of atrophy (left, right) was confirmed using an atrophy lateralization index measured from structural brain MRI, as described in section 2.3.3.3. HC participants who were neurologically normal based on their neurological exam, neuropsychological evaluation and MRI were also included in the sample as a comparison group.

General inclusion criteria for this study were as follows: i) Clinical dementia rating (CDR)≤1 or Mini-Mental State Examination (MMSE) ≥15; (ii) availability of at least the result of one face processing tests; (iii) availability of an MRI scan available within one year of the face processing tests. In total, 74 SD patients fulfilled these criteria. From that sample, we constructed three demographically-(age, sex, education) and clinically-(disease duration, MMSE, CDR total for patients with svPPA and sbvFTD only) matched groups. Our final sample comprised 36 HC, 54 patients with svPPA and 20 with sbvFTD.

### 2.2 Standard protocol approvals, registrations, and patient consent

Written informed consent was obtained from all participants (or legally authorized representatives of participants) in the study and the institutional review board at University of California, San Francisco in the United States approved the study.

### 2.3 Procedure

#### 2.3.1 General neuropsychological assessment

All subjects underwent neuropsychological testing with a comprehensive battery of language, memory, visuospatial, executive functions, and behavior that has been described extensively in Kramer et al. 2003 (Kramer et al., 2003).

#### 2.3.2 Face processing tests

##### 2.3.2.1. Perceptual tests on unknown faces

We used an abridged version of the Benton Facial Recognition Test, comprising items #7 to #13 from the full version (BFRT-S). Participants were simultaneously presented with a target Caucasian face (shown from a frontal viewpoint with a neutral expression) above an array of six test faces with the similar expression. The test contained a total of 7 items and the participant was required to find three images within the six-image array that matched the identity of the target. The six faces in each array varied either in terms of head orientation or lighting. All images were grayscale and displayed the overall shape of the face but were cropped below the chin and beyond the hairline. Participants had an unlimited length of time to complete each trial.

##### 2.3.2.2 Famous faces semantic tests (UCSF Famous Faces Battery)

In the Famous Face Recognition subtest, subjects performed a forced choice task between four faces in which only one was famous. Retrieval of proper name or of semantic/biographical details was not required. Faces were framed with a black oval mask to avoid any possible cueing effects from the pictures’ background. The 20 selected famous faces came from a pool of 200 black-and-white photographs of celebrities in different professional categories whose familiarity was determined by a behavioral study previously described in Gorno-Tempini & Price, 2001(M. Gorno-Tempini et al., 2001). The non-famous faces were matched to the famous ones for mean age, sex and facial expression. All faces were matched for mean luminance.

In the Famous Face Semantic Association pictures (FFSA-P) subtest, subjects were instructed to match two famous faces — among three choices—according to their profession. In each trial, the three famous faces were carefully matched for perceptual characteristics and facial expression. This ensured that inferences based on perceptual similarity alone would not be sufficient to differentiate between the targets and the distractor. Instead, identification of the celebrities and retrieval of semantic/biographical details were necessary to perform the task correctly.

Similarly, in the Famous Face Semantic Association words (FFSA-W) subtest, subjects were instructed to match two famous names — among three choices — according to their profession(M. Gorno-Tempini et al., 2001). Finally, in the Famous Face Confrontation Naming subtest, subjects were prompted to name 20 sequentially presented headshots of celebrities.

#### 2.3.3 Neuroimaging

##### 2.3.3.1. MRI acquisition

MRI data were collected within 1 year of the completing face battery tasks. T1 images were acquired with either a 1.5T (n = 17) (Gorno Tempini et al., 2004), 3T (n = 93) scanner as previously described (Mandelli et al., 2014).

##### 2.3.3.2. MRI processing

Image preprocessing was performed using CAT12, which is a SPM12 (Statistical Parametric Mapping 1) toolbox running on MATLAB (Mathworks, Natick, MA, USA) to conduct VBM. All T1-weighted images were corrected for bias (i.e., field inhomogeneities and noise). They were then segmented into grey matter (GM), white matter and cerebrospinal fluid, spatially normalized to the standard template provided in SPM12, and modulated by the Jacobian determinant to preserve the relative GM volume. After pre-processing, all scans passed a visual check for artifacts and the automated CAT12 quality check protocol. The modulated and normalized GM images were then smoothed with a Gaussian kernel of 8 mm FWHM. Whole-brain statistical maps were first examined at voxel-wise significance level of p < .001 uncorrected. Correction for multiple comparisons was then performed by controlling the family-wise error (FWE) rate at p < .05 at the cluster level.

##### 2.3.3.3. Atrophy predominance characterization

A lateralization index was used to measure the predominance of left or right ATL atrophy based on the calculated GM images as described above. Estimations of left and right ATLs’ volumes were performed using the modulated GM images in SPM12 as the sum of voxel within a mask specific to this region. Z-scores of these volumes were calculated using HC volumes as reference. The lateralization indexes were then extracted subtracting the z-score corresponding to the left and right ATL (left ATL z-score – right ATL z-score). Positive values indicated a right predominant atrophy supporting the diagnosis of sbvFTD, while negative indicated a left predominant atrophy suggestive of imaging-supported diagnoses of svPPA.

### 2.4 Statistical analyses

Data analysis was performed with SPSS (v.29, SPSS/IBM, Chicago, IL, USA). Means of demographic measures, neuropsychological, language, socioemotional and face tasks were compared with the analysis of covariance test correcting for age, sex, education and disease severity as measured by the MMSE.

To determine the GM correlates of perceptual face and semantic task, we used a VBM approach to examine the correlation between each voxel of the whole brain and each task’s performance. We entered the scores for each face processing task in a regression model as a variable of interest, with the normalized, modulated, smoothed GM images as inputs, and including age, sex, number of days between the MRI and intracranial volume as covariates of no interest. Contrasts were set to examine the hypothesis that a lower score on face tasks would be associated with decreased GM volume. These association analyses were conducted on both svPPA and sbvFTD patients combined. The statistical threshold was set at p < 0.05 FWE cluster corrected.

## 3. Results

### 3.1 Characterization of HC, svPPA, and sbvFTD participants

The sample comprises three demographically-(age, sex, education) and clinically-(disease duration, clinical dementia rating (CDR) total, CDR sum of boxes score for patients with svPPA and sbvFTD only) matched groups. Summary statistics for demographic characteristics, and cognitive and language testing for all three groups are presented in Table 1. As expected, patients with svPPA had significantly lower scores than patients with sbvFTD in verbal semantics tests (naming, semantic and phonemic fluency, word-picture matching, irregular words reading) and patients with sbvFTD had significantly lower informer-rated empathy score (RSMS), and visual episodic memory (Modified Rey Recognition) than patients with svPPA.

**Table 1.**
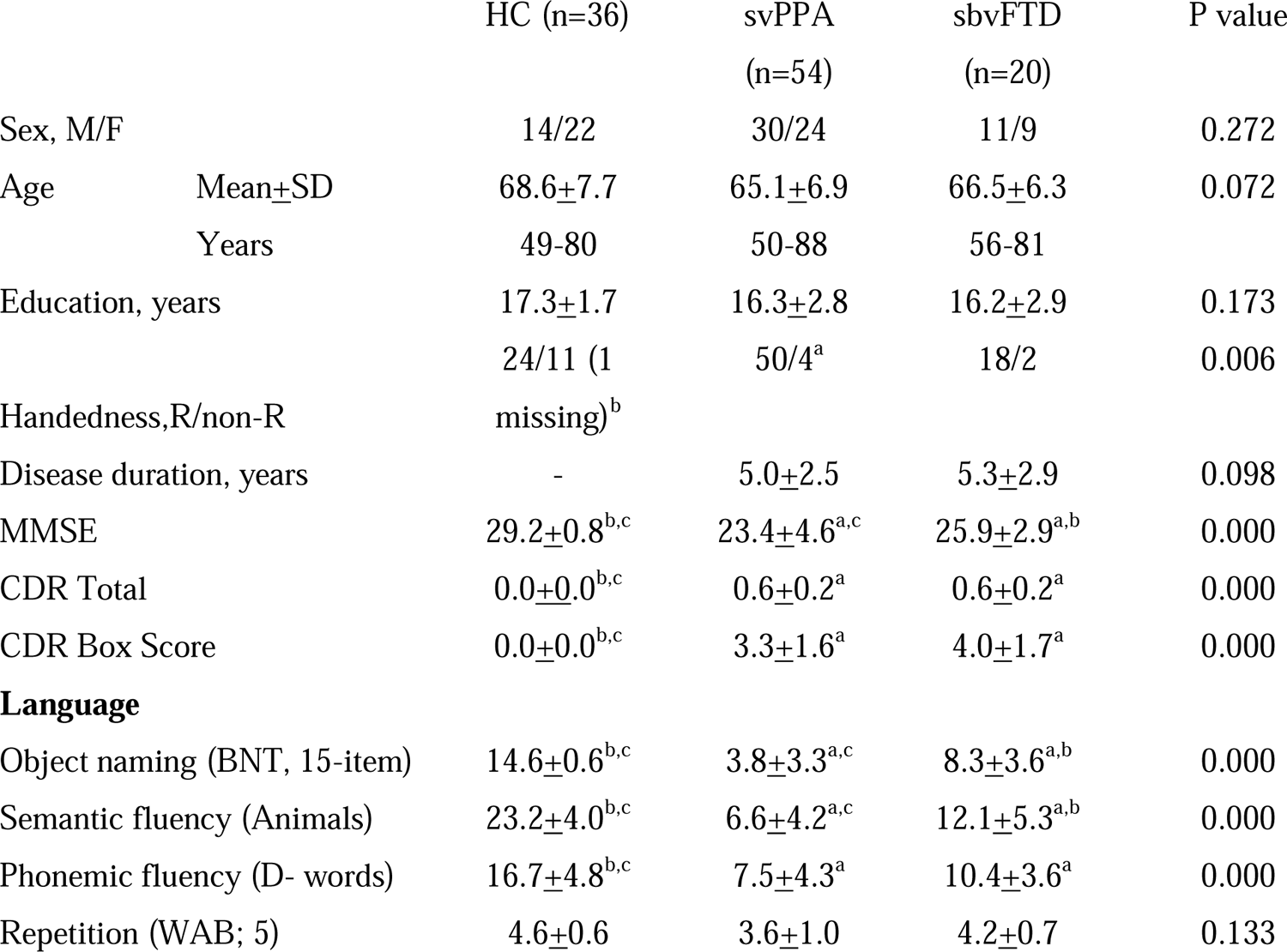
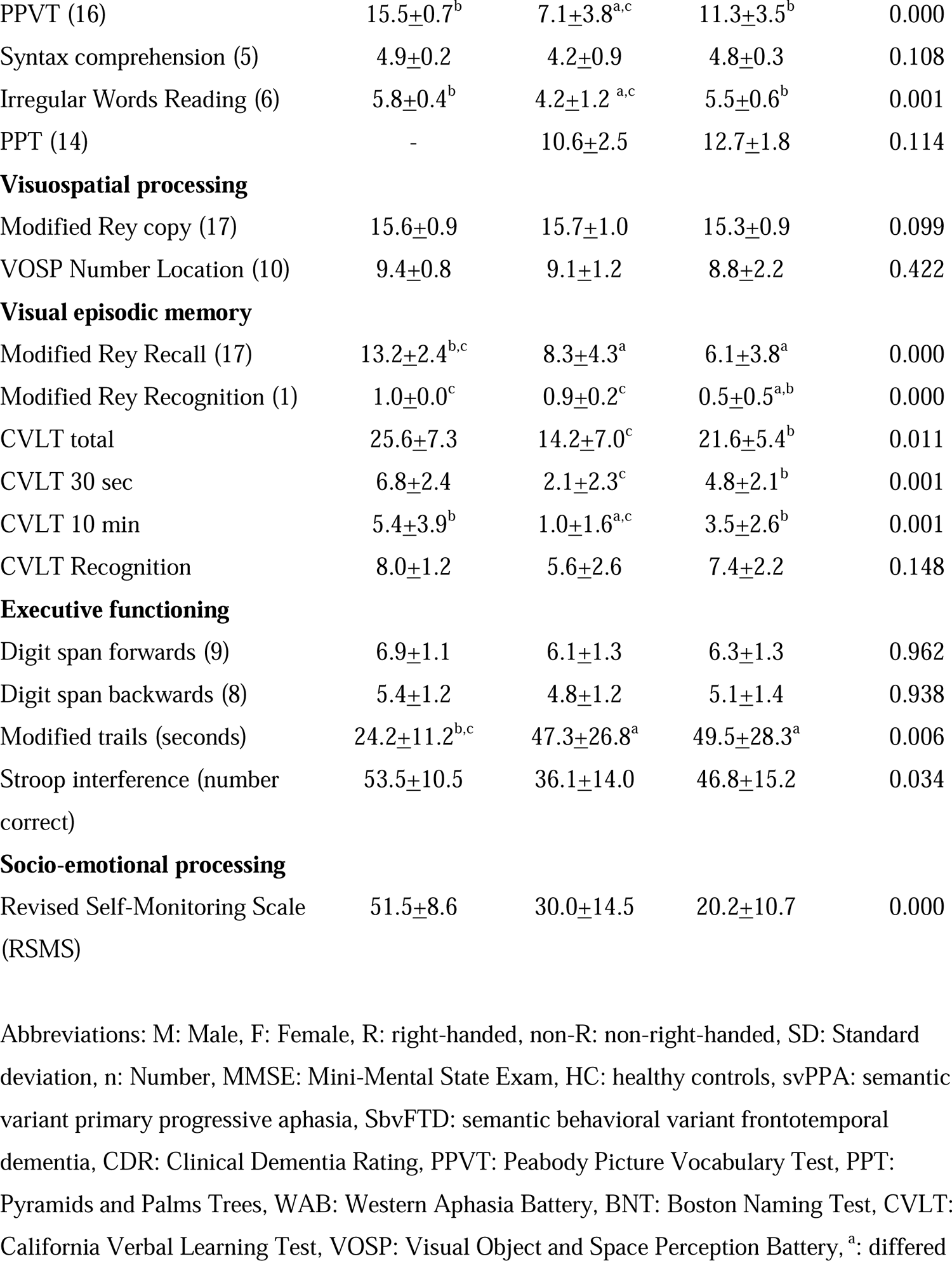
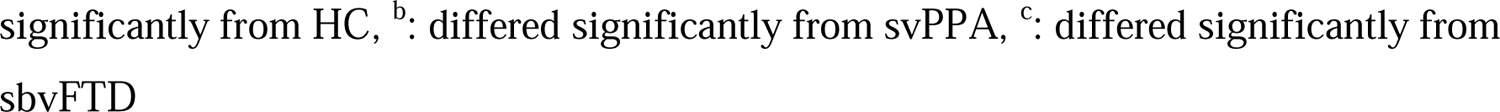
Demographics, neuropsychological and language data for all groups.

### 3.2 Characterization of HC, svPPA, and sbvFTD participants’ performance on face processing tests

Face processing tasks comparisons were performed between the three groups (Table 2, Figure 1). In terms of perceptual face processing, no statistical significance was observed between the three groups on the BFRT-S. In terms of semantic face processing, both patients with svPPA and sbvFTD performed significantly lower on all semantic face tasks in comparison to HC. SbvFTD had significantly lower scores on famous face recognition subtest compared to svPPA. Conversely, svPPA patients performed significantly worse on famous face confrontation naming compared to sbvFTD patients. On both FFSA-P and FFSA-W tasks, svPPA and sbvFTD patients had a similar performance when compared to each other.

**Figure 1:**
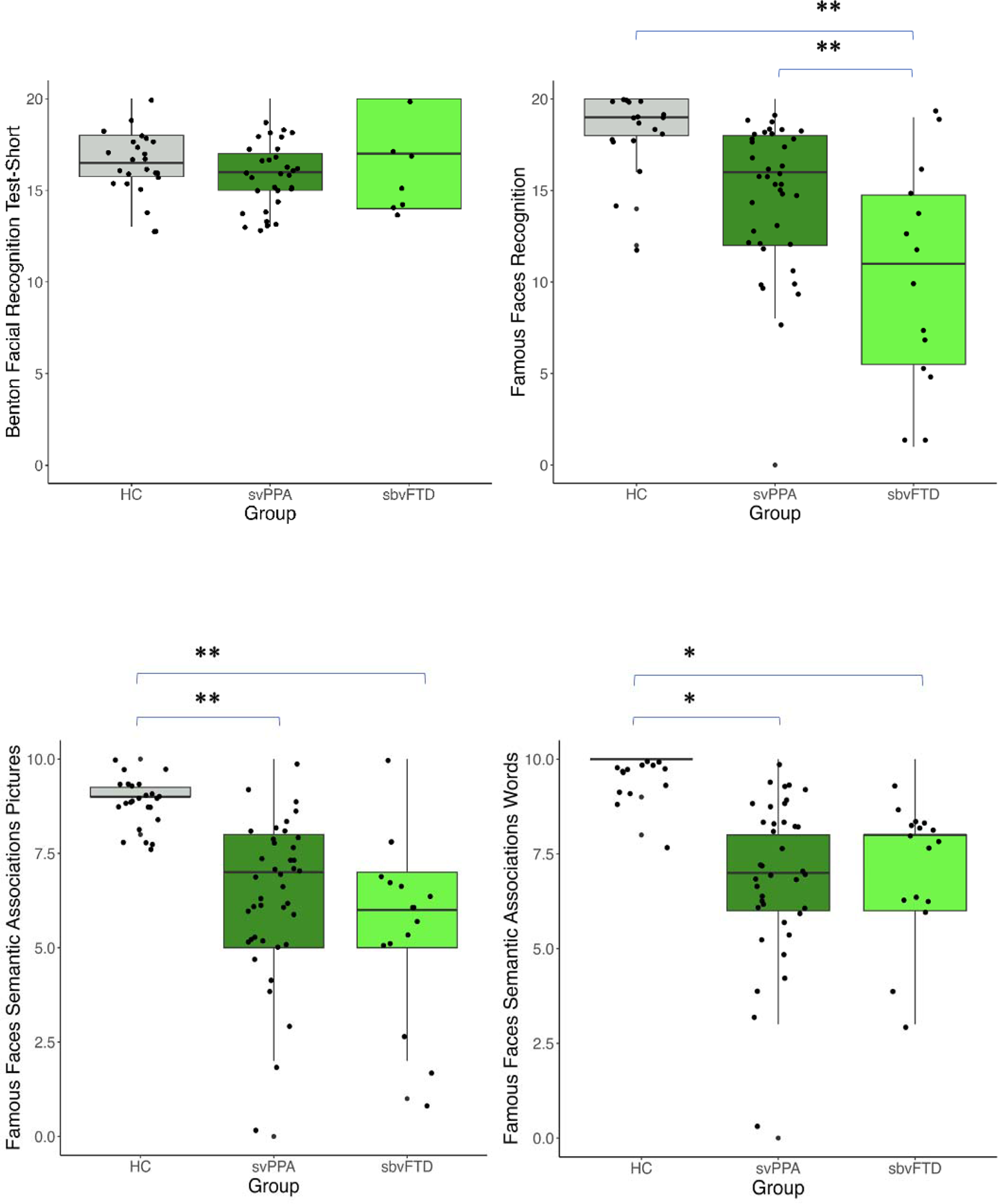
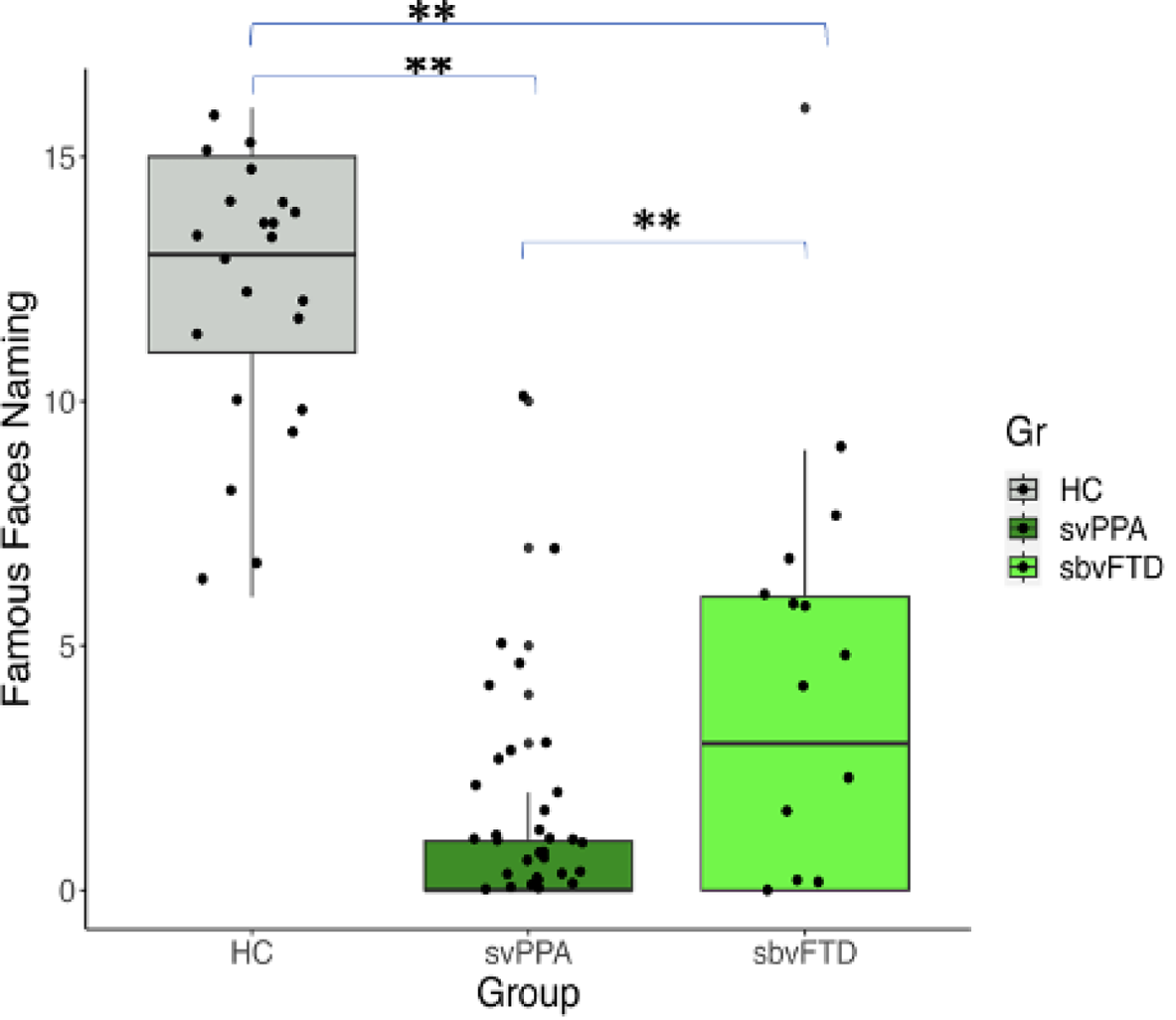
Perceptual face, semantic face and emotional facial expression recognition tasks breakdown among SD patients and HC. The results of the face perception, and the four tasks of the UCSF famous faces battery allow descriptive comparisons of performance on these tasks between SD patients and HC (adjusted for age, sex, education, and MMSE) Abbreviations: HC: healthy controls, svPPA: semantic variant primary progressive aphasia, sbvFTD, semantic behavioral variant frontotemporal dementia

**Table 2.**
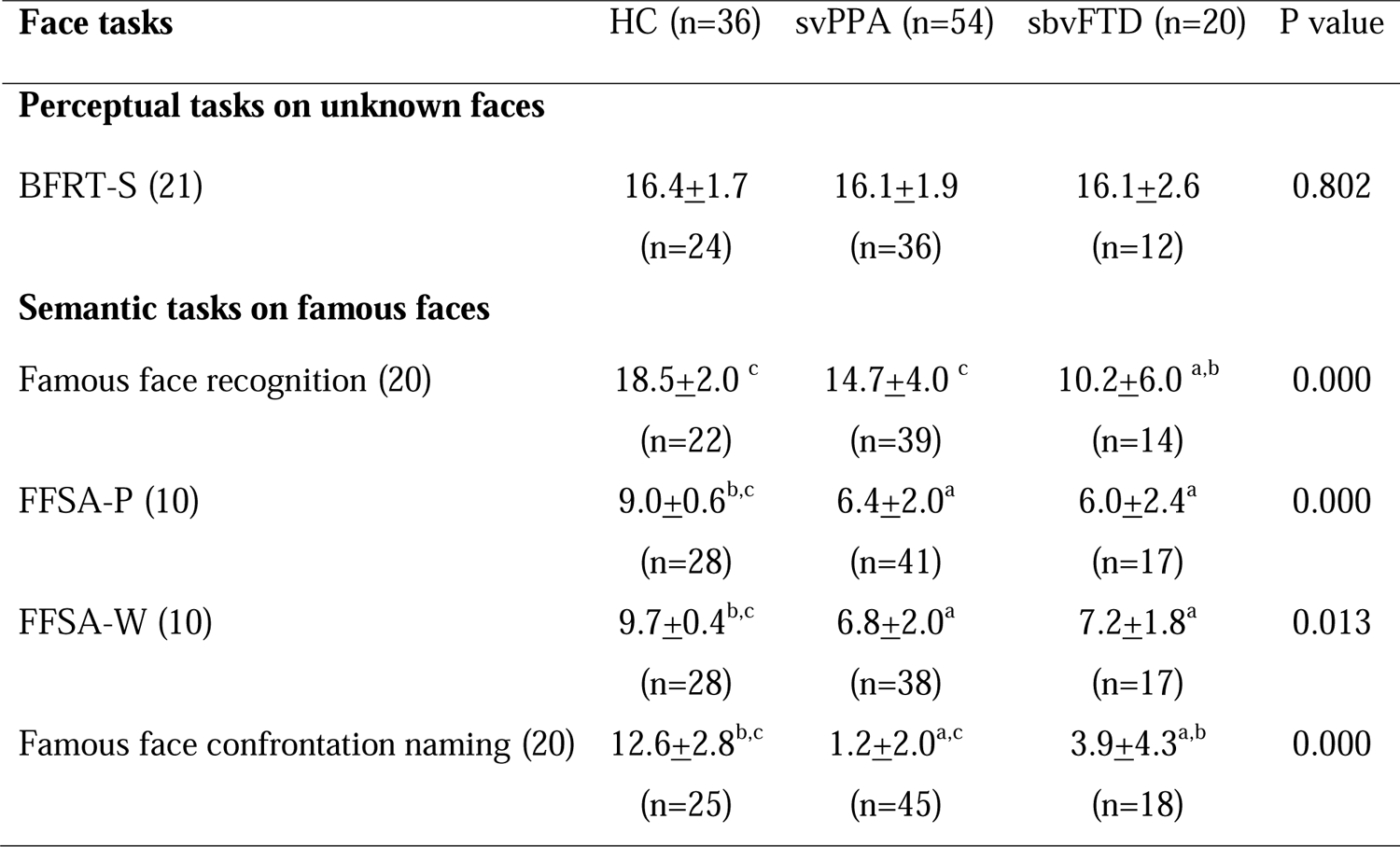
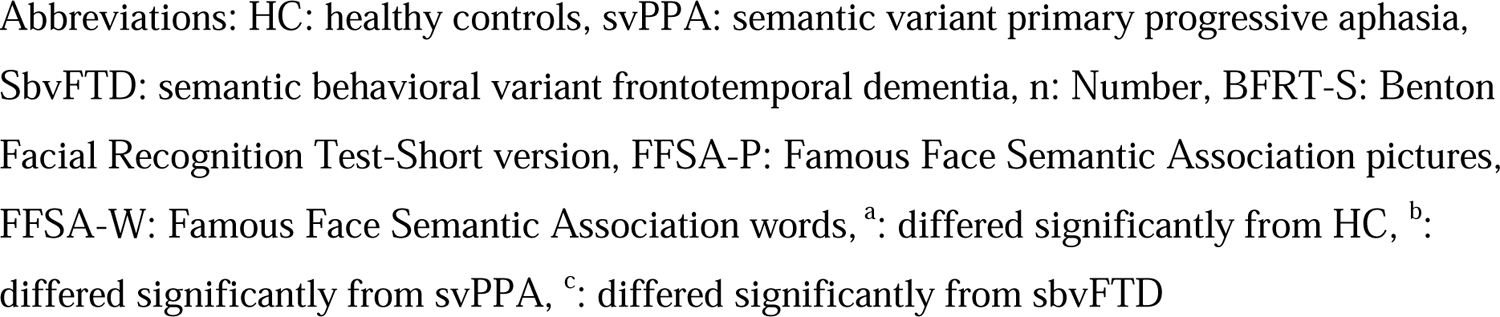
Comparing the performance of HC and SD patients on different face battery tasks (adjusted for age, sex, education, MMSE)

| Face tasks | HC (n=36) | svPPA (n=54) | sbvFTD (n=20) | P value |
| --- | --- | --- | --- | --- |
| **Perceptual tasks on unknown faces** |  |  |  |  |
| BFRT-S (21) | 16.4±1.7<br>(n=24) | 16.1±1.9<br>(n=36) | 16.1±2.6<br>(n=12) | 0.802 |
| **Semantic tasks on famous faces** |  |  |  |  |
| Famous face recognition (20) | 18.5±2.0 <sup>c</sup><br>(n=22) | 14.7±4.0 <sup>c</sup><br>(n=39) | 10.2±6.0 <sup>a,b</sup><br>(n=14) | 0.000 |
| FFSA-P (10) | 9.0±0.6 <sup>b,c</sup><br>(n=28) | 6.4±2.0 <sup>a</sup><br>(n=41) | 6.0±2.4 <sup>a</sup><br>(n=17) | 0.000 |
| FFSA-W (10) | 9.7±0.4 <sup>b,c</sup><br>(n=28) | 6.8±2.0 <sup>a</sup><br>(n=38) | 7.2±1.8 <sup>a</sup><br>(n=17) | 0.013 |
| Famous face confrontation naming (20) | 12.6±2.8 <sup>b,c</sup><br>(n=25) | 1.2±2.0 <sup>a,c</sup><br>(n=45) | 3.9±4.3 <sup>a,b</sup><br>(n=18) | 0.000 |

Abbreviations: HC: healthy controls, svPPA: semantic variant primary progressive aphasia, SbvFTD: semantic behavioral variant frontotemporal dementia, n: Number, BFRT-S: Benton Facial Recognition Test-Short version, FFSA-P: Famous Face Semantic Association pictures, FFSA-W: Famous Face Semantic Association words, <sup>a</sup>: differed significantly from HC, <sup>b</sup>: differed significantly from svPPA, <sup>c</sup>: differed significantly from sbvFTD

### 3.3 Neural correlates of face processing tests in SD patients

In terms of perceptual face processing, the regression voxel-based analysis (controlling for age, sex, and total intracranial volume) showed a correlation between BFRT-S and GM volume in a cluster centered in right pSTS (Figure 2) (Table 3).

**Figure 2:**
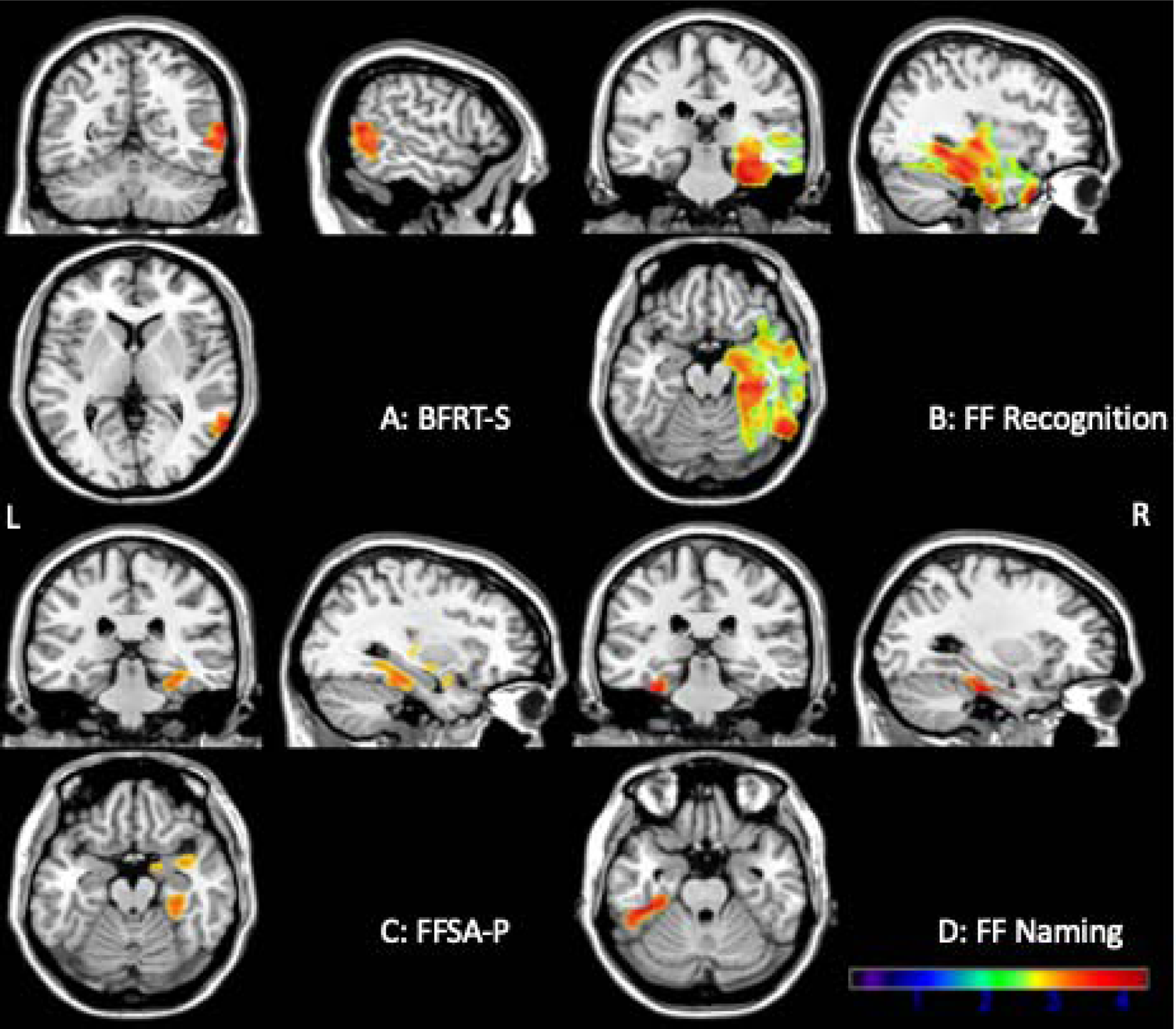
GM correlates of perceptual, semantic face and emotional facial expression recognition tasks in svPPA and sbvFTD patients adjusted for age, sex and total intracranial volume (p > .05 FWE cluster-corrected).

**Table 3.**
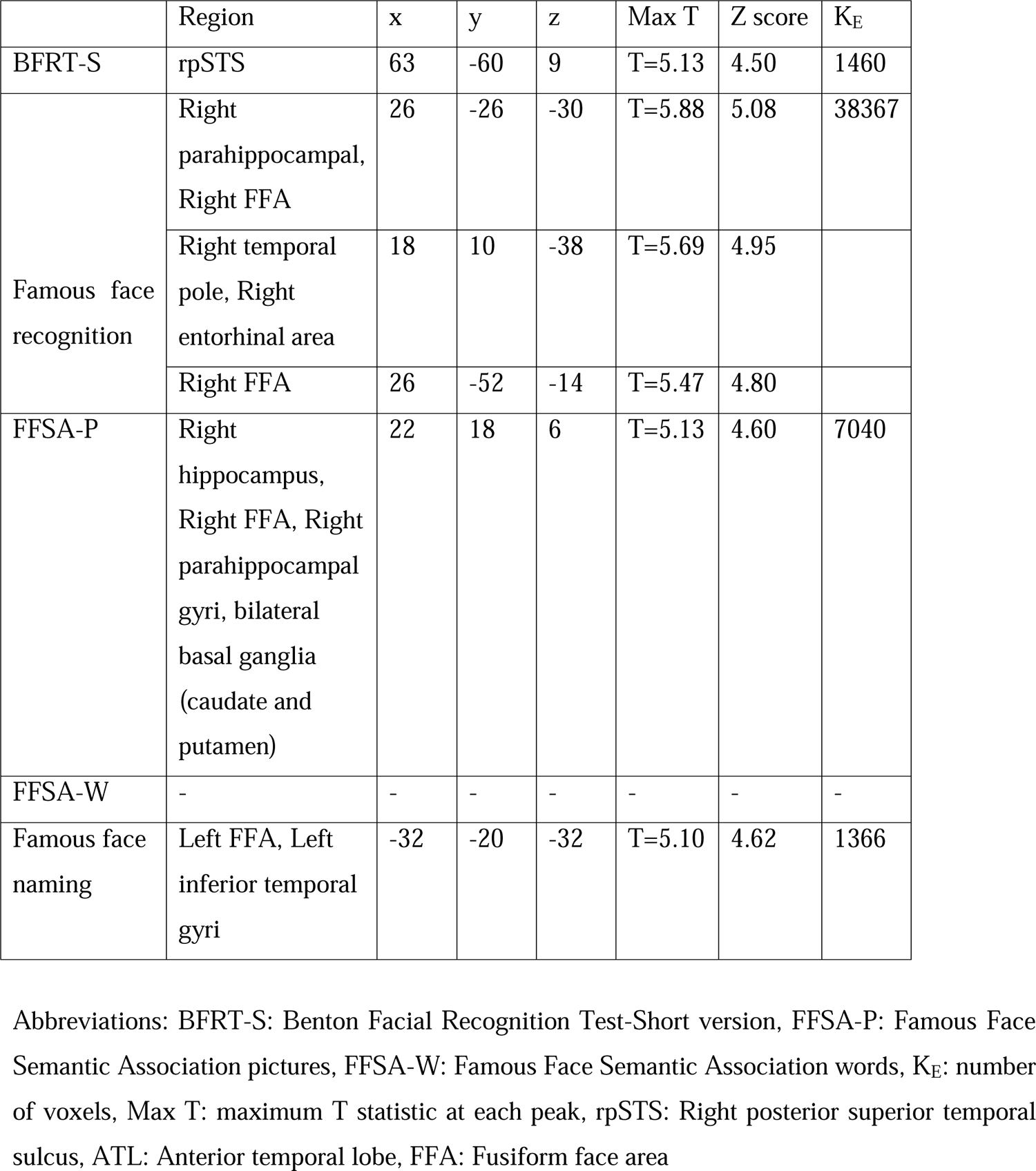
Coordinates of voxel-based morphometry analysis of face tasks (p > .05 FWE cluster-corrected)

|  | Region | x | y | z | Max T | Z score | K<sub>E</sub> |
| --- | --- | --- | --- | --- | --- | --- | --- |
| BFRT-S | rpSTS | 63 | -60 | 9 | T=5.13 | 4.50 | 1460 |
| Famous face recognition | Right parahippocampal, Right FFA | 26 | -26 | -30 | T=5.88 | 5.08 | 38367 |
|  | Right temporal pole, Right entorhinal area | 18 | 10 | -38 | T=5.69 | 4.95 |  |
|  | Right FFA | 26 | -52 | -14 | T=5.47 | 4.80 |  |
| FFSA-P | Right hippocampus, Right FFA, Right parahippocampal gyri, bilateral basal ganglia (caudate and putamen) | 22 | 18 | 6 | T=5.13 | 4.60 | 7040 |
| FFSA-W | - | - | - | - | - | - | - |
| Famous face naming | Left FFA, Left inferior temporal gyri | -32 | -20 | -32 | T=5.10 | 4.62 | 1366 |

Abbreviations: BFRT-S: Benton Facial Recognition Test-Short version, FFSA-P: Famous Face Semantic Association pictures, FFSA-W: Famous Face Semantic Association words, K<sub>E</sub>: number of voxels, Max T: maximum T statistic at each peak, rpSTS: Right posterior superior temporal sulcus, ATL: Anterior temporal lobe, FFA: Fusiform face area

In terms of semantic face processing in SD patients, famous face recognition subtest scores correlated with GM volumes in the right parahippocampal and FFA. Similarly, on the visual semantic association task, FFSA-P scores correlated with a cluster centered in hippocampus, FFA, and parahippocampal gyri on the right, and extending into the bilateral basal ganglia, namely caudate and putamen. SD patients’ scores on verbal semantic association task, FFSA-W, showed no correlation at the cluster level at FWE-corrected at p < .05 FWE cluster-level. However, at voxel-wise significance level of p < .001 uncorrected, FFSA-W showed a correlation with left ATL, left frontal operculum and left insula. Finally, scores on famous face confrontation naming showed a significant correlation with volumes in the left FFA and left inferior temporal gyri.

### 3.4 Post-hoc analysis

Observing a correlation between BFRT-S and right pSTS, we conducted supplementary analyses to see if the above correlation was driven by SD patients with higher disease severity. First, we assessed the correlations between BFRT-S and two measures of disease severity, namely the Mini-Mental State Exam (MMSE) and CDR Sum of Boxes score. We found a non-significant statistical trend towards a positive correlation between the BFRT-S and MMSE (r = 0.282; p = 0.067) and a non-significant statistical trend towards a negative correlation between the BFRT-S and CDR (r=-0.232; p = 0.113), suggesting that patients with higher disease severity show more severe perceptual face processing deficits on the BFRT-S (Figure 3). Second, we also re-ran the VBM analysis investigating the neural correlates of the BFRT-S including the CDR as a covariate, to verify whether doing this would remove the statistically significant cluster in the rpSTS. Indeed, the correlation between BFRT-S and GM volume in the right pSTS became insignificant when controlling for CDR.

**Figure 3:**
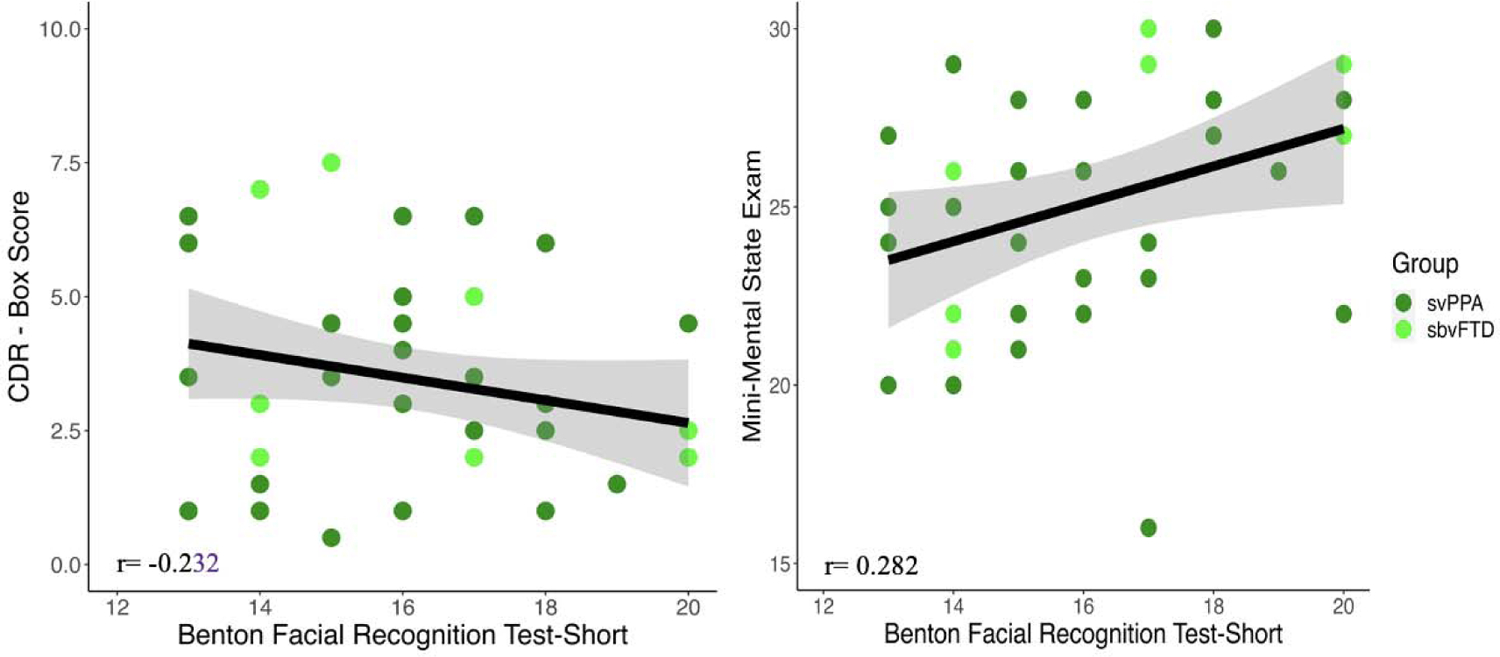
A: Correlation between BFRT-S and CDR box score in svPPA and sbVFTD groups, B: Correlation between BFRT-S and Mini-mental status exam in svPPA and sbVFTD groups Abbreviations: svPPA: semantic variant primary progressive aphasia, sbvFTD: semantic behavioral variant frontotemporal dementia, CDR: Clinical Dementia Rating

## 4. Discussion

This study primarily aims to determine if the impaired recognition of familiar faces observed in SD patients can be solely attributed to a semantic deficit, or if a perceptual face deficit also contributes to this symptom. We found that SD patients demonstrated widespread semantic deficits in famous face recognition and naming compared to controls, with sbvFTD patients performing worse than svPPA patients in famous face recognition, and svPPA patients showing a more impaired performance in famous face naming compared to sbvFTD patients. Both SD groups also showed deficits in verbal and visual semantic faces association tasks in comparison to HC. Regarding perceptual face processing, svPPA and sbvFTD patients showed no significant difference in performance in comparison to HC. The current study also informs on the neural bases of face processing, with perceptual processes being associated with GM volume in the right pSTS and semantic processes being mostly associated with GM volume in the right and/or left ATL, depending on the visual or verbal predominance of the task.

### 4.1. Face processing in SD: from perception to semantics

Our findings provide empirical evidence of the cognitive and neuroanatomical decoupling between perceptual and semantic face processing steps. First, SD patients (both svPPA and sbvFTD) did not show any impairment in the perceptual face processing test, where the participant needs to have the ability to select the three similar faces with a different head orientation and lightening to the target face among six different faces. In line with our study, Kamminga et al. did not observe impaired face perception when they compared right SD patients to bvFTD and HC (Kamminga et al., 2015). However, our results are partially inconsistent with two studies demonstrating impairments on a face perception task in SD, in both right- and left-SD patients (Ding et al., 2020) or in right SD only (Kumfor et al., 2015). These conflicting results might be due to differences in the demographic/clinical characteristics of the samples. In the current study, we restricted the sample to SD cases with a clinical dementia rating (CDR) score of ≤1 or a Mini-Mental State Examination (MMSE) of ≥15 to exclude severe cases, and our sample was also largely more educated (approximately 5 more years of education on average). Alternatively, the inconsistency might be due to the different perceptual facial processing tests used. Our posthoc analysis showed trends toward a significant correlation between disease severity and face processing. Furthermore, the significant correlation between BFRT-S and the GM volume in the rpSTS was mainly driven by disease severity. This suggests that the most severe SD patients from our sample are beginning to show perceptual face processing deficit. Therefore, taking all evidence together, we suggest that there are no perceptual face processing deficits in the early stages of SD, but that these might appear later in the disease especially in those with right posterior temporal atrophy.

In terms of semantic face tasks, although sbvFTD patients showed difficulty with all semantic face tasks, they particularly had an impaired performance in famous face recognition compared to svPPA patients. Instead, svPPA patients had poor scores in famous face naming despite their spared performance on familiarity judgment task. This aligns with the clinical description of sbvFTD syndrome in which a loss of person-specific semantic knowledge is reported as one of the early core symptoms(Younes et al., 2022). Consistent with previous studies, these findings suggest that these two tests can be particularly useful as a clinical marker to support the diagnosis of sbvFTD, as well as for the differential diagnosis with svPPA(Binney et al., 2016; Borghesani et al., 2019; Irish et al., 2013; Luzzi et al., 2017; Pozueta et al., 2019) and should therefore be prioritized in clinical practice. This result could also be in line with the visual (famous face recognition) or verbal (famous face naming) predominance of the task, as the right ATL is thought to be particularly involved in visual semantics and the left ATL in verbal semantics (Binney et al., 2016; Borghesani et al., 2019).

In light of these results, we can conclude that the face processing deficits experienced by SD patients is different than the condition called prosopagnosia, as defined by the incapacity to identify familiar persons on the basis of visual perception of their faces only. However, the semantic facial processing deficit seen in SD patients is multimodal which is different from the unimodal visual facial perceptual deficit seen in prosopagnosia. In our study, SD patients demonstrated an impairment in verbal semantic task (i.e, FFSA-W) in addition to impairment in famous face recognition and visual semantic task. Moreover, another previous study showed that subjects with bilateral anterior temporal lesions had deficits in the recognition of voices and not only faces (Liu et al., 2016). As a result, the use of the term prosopagnosia can be imprecise or misleading as SD patients have person-related multimodal semantic deficits(Liu et al., 2016).

Future studies shall aim to integrate more non-verbal auditory inputs (i.e., famous people voices), while comparing performance across tasks explicitly addressing different cognitive processes, as done here. This will be helpful in understanding the differences in processing the visual vs auditory stimuli with regards to perceptual and semantic cognitive tasks.

### 4.2 Anatomical model of face processing

In our study, we found that face perception is correlated with right pSTS in SD patients, a region described as part of the core system of face processing (Haxby et al., 2000). This is in line with the previous studies (D. I. Perrett et al., 1990; D. I. Perrett et al., 1984; Pitcher, 2014; Pitcher et al., 2011) who have also found cells in the superior temporal sulcus that respond selectively to different individuals and expressions. They reported that the clusters of cells in the superior temporal sulcus that respond to different aspects of faces are intermixed with clusters of cells that respond to other visual features, most notably movement of the face, head and body (Oram et al., 1996; D. Perrett et al., 1985). Moreover, pSTS has also been variably associated with many functions, including the perception of biological motion (Allison et al., 2000), and coding of specific gaze directions and expressions (Calder et al., 2007; Said et al., 2010; Winston et al., 2004). This could explain the correlation between right pSTS and the BFRT-S test since the faces tested had different head orientation, which could explain the significant correlation identified with the right pSTS volume. This region is not usually damaged in SD, at least in the first stages of the disease, consistent with the results of our study.

With regards to the extended system of face processing (Haxby et al., 2000), and in line with the previous studies (Binney et al., 2016; Borghesani et al., 2019; Gainotti, 2007), we have found that damage to the right FFA, hippocampus, parahippocampus, and entorhinal areas on the right were associated with semantic face processing (impaired performance on familiarity judgment and visual semantic association task). On the other hand, ATL, inferior temporal and FFA on the left were correlated with the verbal semantic association task and famous face naming. In a previous study, the neural response in the FFA was shown to be consistently associated with changes in face identity specifically, suggesting that this region is not sensitive to subtle physical changes and thus not involved in face perception (Ramon et al., 2010). The differential functional specialization of the left and right ATL based on type of semantic features and task supports the model that the concerted functionality of both hemispheres is required for the successful identification of famous people (Gainotti, 2015a, 2015b; Ralph et al., 2017). In our study, we also found a lateralized correlations between verbal tasks such as FFSA-W and naming with left ATL. Moreover, we showed that right ATL correlated with the FFSA-P and famous face recognition tasks.

### 4.4. Limitations

First, while the five face processing tasks were carefully designed to separately assess key processing steps and to avoid perceptual confounds, tasks involving famous faces are always challenging to build to avoid biases. For example, the famous faces used in these tasks mostly belong to 19s, the younger patients could have had difficulty recognizing some of the faces. Although we have found that SD patients showed impairment in both verbal and visual semantic association tasks as shown on FFSA-P and FFSA-W, our results cannot be generalized to famous person identification as achieved through other sensory modalities (Gainotti, 2015a, 2015b). Moreover, since sbvFTD and svPPA progress into similar clinical profiles as atrophy spreads, it may be interesting to longitudinally assess the changes in face processing in patients with SD (Borghesani et al., 2019). On the imaging side, while we have found the different temporal brain regions to be correlated with the face processing tasks, it would be interesting to use connectivity-based imaging approaches in a future study to investigate the possible larger and bilateral brain network involved in face processing in SD patients. Finally, we acknowledge the serious limitation that patients included in this study were mostly white, highly educated and native English speakers. Further studies that include more diverse patient populations are needed to shed light on the cultural and environmental variability of face processing in patients with SD.

### 4.5. Conclusion

In conclusion, we found that SD patients did not suffer from perceptual face processing deficit at an early stage in the disease course. Moreover, GM volume in rpSTS was correlated with the face perception abilities. In sbvFTD patients, a loss of person-specific knowledge was seen in relation to right ATL-predominant degeneration and decline in the neural networks that support non-verbal, socioemotional semantic knowledge. On the other hand, svPPA patients demonstrated an impaired performance on the verbal semantic knowledge task mainly related to left ATL-predominant atrophy. Specific neuropsychological tests that investigate face perceptual and semantic processing are important for capturing early sbvFTD symptoms and should be included in standard evaluations to help with better prognostication and therapeutics.

## Data availability

Public archiving is not yet permitted under the study’s IRB approval due to the sensitive nature of patient data, although we can share anonymized data. Specific requests can be submitted through the UCSF-MAC Resource (Request form: http://memory.ucsf.edu/resources/data). Following a UCSF-regulated procedure, access will be granted to designated individuals in line with ethical guidelines on the reuse of sensitive data. This would require submission of a Material Transfer Agreement, available at: https://icd.ucsf.edu/material-transfer-and-data-agreements. Commercial use will not be approved.

## Acknowledgements

The study was supported by grants NINDS R01 NS050915, NIA P50 AG023501, NIA P01 AG019724, NIDCD K24 DC015544, NIA U01 AG052943, NIA R01 AG038791, NIA U54 NS092089, NIA U01 AG045390, K23DC018021. Additional funds include the Larry Hillblom Foundation and the Global Brain Health Institute. These supporting sources had no involvement in the study design, collection, analysis or interpretation of data, nor were they involved in writing the paper or the decision to submit this report for publication. The authors thank the patients and their families for the time and effort they dedicated to the research.

## 4.7. Author contributions

Golnaz Yadollahikhales: Conceptualization, Methodology, Formal analysis; Maria Luisa Mandelli: Software, formal analysis; Zoe Ezzes: Resources; Janhavi Pillai: Software; Buddhika Ratnasiri: Resources; David Paul Baquirin: Resources; Zachary Miller: Investigation, Resources; Jessica de Leon: Investigation, Resources; Boon Lead Tee: Resources, Investigation; William Seeley: Resources; Howard Rosen: Resources; Bruce Miller: Resources; Joel Kramer: Resources; Virginia Sturm: Resources, Investigation; Maria Luisa Gorno-Tempini: Conceptualization, Methodology; Maxime Montembeault: Conceptualization, Methodology

